# Irritability in young people with copy number variants associated with neurodevelopmental disorders (ND-CNVs)

**DOI:** 10.1101/2023.12.05.23299440

**Authors:** Jessica H. Hall, Samuel. J.R.A Chawner, IMAGINE-ID consortium, Jeanne Wolstencroft, David Skuse, Peter Holmans, Michael J. Owen, Marianne B.M. van den Bree

## Abstract

**Background:** A range of rare mutations involving micro-deletion or -duplication of genetic material (copy number variants (CNVs)) have been associated with high neurodevelopmental and psychiatric risk (ND-CNVs). Irritability is frequently observed in childhood neurodevelopmental conditions, yet its aetiology is largely unknown. Genetic variation may play a role, but there is a sparsity of studies investigating presentation of irritability in young people with ND-CNVs.

**Aims:** This study aimed to investigate whether there is a difference in irritability in young people with rare ND-CNVs compared to those without ND-CNVs, and to what extent irritability is associated with psychiatric diagnoses and cognitive ability (IQ).

**Methods:** Irritability and broader psychopathology was assessed in 485 young people with ND-CNVs and 164 sibling controls, using the child and adolescent psychiatric assessment (CAPA). Autism was assessed using the Social Communication Questionnaire (SCQ), and Intelligence Quotient (IQ) by the Wechsler Abbreviated Scale of Intelligence (WASI).

**Results:** 54% of young people with ND-CNVs met the threshold for irritability; significantly more than controls (OR = 3.77, CI = 3.07-7.90, p= 5.31 x 10^-11^). When controlling for the presence of other psychiatric comorbidities, ND-CNV status was still associated with irritability. There was no evidence for a relationship between irritability and IQ.

**Conclusions:** Irritability is an important aspect of the clinical picture in young people with ND-CNVs. This work shows that genetic variation is associated with irritability in young people with ND-CNVs, independent of psychiatric comorbidities or IQ impairment. Clinicians should be aware of this increased risk to inform management and interventions.

## Introduction

### Neurodevelopmental Risk Copy Number Variants (ND-CNVs)

Copy number variation (CNV) refers to variation in the genome, in which an individual’s number of copies of a specific segment of DNA is altered^1^. Commonly, these structural differences come about through the duplication or deletion of genetic material^1^. A number of CNVs show associations with increased risk of neurodevelopmental disorders (ND-CNVs)^2^, including attention deficit hyperactivity disorder (ADHD)^3^, autism and intellectual disability^4-7^, as well as other mental health conditions such as schizophrenia, and mood and anxiety disorder^8^. Whilst ND-CNVs are individually rare, they are collectively more common, with population based studies estimating that ∼1 in 200 newborns has either a deletion or duplication in neurodevelopmentally associated regions^9^. With the improvement of genomic technology, ND-CNVs are being increasingly recognised, and rates of diagnosis can be expected to continue to rise; they are thus of increasing relevance to clinical practice.

### Chronic Irritability

Severe irritability is a common presentation in those referred to mental health services as children and adolescents^10^. Irritability can be clinically defined as an increased proneness to anger relative to peers at the same developmental level^10^. In clinical literature the terms anger, rage and frustration tend to be used interchangeably with “irritability”, but all refer to developmentally inappropriate expressions of the emotion of irritability, or dysregulation^11^. Currently, irritability is often conceptualised as a distinct homogenous construct within psychiatric phenomenology, but it is also recognised as a core or accompanying feature of several psychiatric disorders^10,11^. Chronic irritability is described in the literature as being present if a child is prone to feelings of anger, bad temper, or resentment at least 45 times in the past 3 months (i.e., approximately 3–4 times/week)^12^. The impact of chronic irritability on the individual and their family members makes it an important symptom to address clinically.

### Irritability, Psychopathology and Cognition

Often defined as a mood, irritability is present in the DSM-V and ICD-10 criteria of bipolar disorder and depression, and is often observed in children with oppositional defiant disorder (ODD) and conduct disorder (CD)^13^. Whilst not a defining diagnostic criterion, irritability is also an especially common feature of ADHD; with an estimated prevalence of 91%, and there is evidence of clinical and genetic overlap between irritability and ADHD^14,15^. This has led to the suggestion that irritability should perhaps be conceptualised as a neurodevelopmental problem, rather than a mood problem in children^15^. Children with irritability are also more likely to experience comorbid anxiety^16^, and longitudinal studies have demonstrated associations between early irritability and later depression and anxiety^16-18^. Irritability is also recognised as a prevalent and impairing problem in children with autistic spectrum disorders (ASD)^10,11,19^. Furthermore, irritability has also been associated with impaired cognitive functioning^20^.

### Irritability in ND-CNVs

Despite irritability being a prominent feature of many of the psychiatric and neurodevelopmental diagnoses seen in young people with ND-CNVs, few studies have investigated the presentation of irritability in this high-risk population subgroup. Research designs using a genotype-first approach whereby young people with ND-CNVs are identified to take part in deep-phenotyping assessments, provide an opportunity to investigate irritability in a population of individuals with a known genetic aetiology, and can contribute novel insights into the links between irritability and mental health conditions. Given that youth with severe irritability display impaired cognitive flexibility compared to youth without irritability and that the presence of ND-CNVs is associated with cognitive deficits^6,8,21^ it could be hypothesised that reduced cognitive ability may contribute to irritability in young people with ND-CNVs.

Better understanding the phenotype of this group of vulnerable young people will help with the development of tailored interventions. This work therefore aims to reduce some of the gaps in the literature by: 1) Investigating whether young people with ND-CNVs are more irritable compared to siblings without known rare variants (controls); 2) Ascertaining whether shared genetic and environmental factors have an effect on the irritability outcome in young people 3) Exploring to what extent ND-CNV status is associated with irritability when controlling for psychiatric diagnoses 3) Studying to what extent ND-CNV status is associated with irritability when controlling for cognitive ability.

## Methodology

### Sample

Families of young people with ND-CNVs were recruited by the The Cardiff rarE genetiC variant researcH prOgramme (ECHO). (https://www.cardiff.ac.uk/centre-neuropsychiatric-genetics-genomics/research/themes/developmental-psychiatry/copy-number-variant-research-group) and IMAGINE-ID (https://imagine-id.org/) studies through Medical Genetics clinics across the UK, charities for chromosomal conditions (Unique, MaxAppeal, 22Crew) and word of mouth.

Participants were recruited on the basis of having a ND-CNV of interest (genotype-first approach) (n=485), and not on the presence of a psychiatric phenotype (phenotype-first approach). Supplementary Table 1 illustrates sample sizes for each ND-CNV. ND-CNV was defined as genetic variants which were a) recurrent (i.e the same mutation occurring at the same location of the genome, in multiple people in a population), and pathogenic or likely pathogenic variants, according to the American College of Medical Genetics and Genomics guidelines^22^ b) associated with neurodevelopmental outcomes^5^. A sibling with no ND-CNV (control) and closest in age to the index child (proband) was invited to take part (n=164). Sample demographic information is presented in Table 2. Written consent was given by the primary caregiver and consent or assent by the young people as applicable. The authors assert that all procedures contributing to this work comply with the ethical standards of the relevant national and institutional committees on human experimentation and with the Helsinki Declaration of 1975, as revised in 2008. All procedures involving human subjects/patients were approved by the appropriate university and National Health Service (NHS) ethics and research and development committees; the NHS London Queen Square research ethics committee (14/LO/1069) and South East Wales Research Ethics Committee (09/WSE04/22). The presence of ND-CNVs in the ND-CNV group, and the absence of the ND-CNV in the control group were confirmed by NHS Medical Genetics Laboratories records, and subsequently, the laboratory of the Cardiff University Division for Psychological Medicine and Clinical Neurosciences (CU DPMCN), using microarray techniques.

### Measures

#### Assessment of irritability, neuropsychiatric and cognitive phenotypes

Assessments were carried out by experienced research psychologists. Participants were given the choice of clinic or in-home assessments, with a view to reducing bias or discrimination against those who may struggle to travel to a research facility; the majority opted for home visits. During assessments, detailed demographic information was collected, along with in-depth evaluation of developmental, behavioural, cognitive and psychiatric features. Information about psychotropic medication use is listed in Supplementary Table 2.

#### Irritability and Psychopathology

The Child and Adolescent Psychiatric Assessment (CAPA) is an in-depth, semi-structured psychiatric interview that allows for establishment of DSM psychiatric diagnoses^23^. The CAPA was carried out by trained research psychologists with the primary caregiver. Interviews were audiotaped and a research diagnosis based on DSM-V, was obtained through consensus meetings led by a child and adolescent psychiatrist. We used the CAPA to establish research diagnoses of attention deficit hyperactivity disorder (ADHD), anxiety and mood disorder (depression, mania or hypomania). Anxiety included any diagnosis of generalised anxiety disorder, social phobia, specific phobia, separation anxiety, panic disorder with and without agoraphobia, and panic disorder. Autism symptomatology was measured by the Social Communication Questionnaire (SCQ)^24^. A cutoff of >15 was deemed to be indicative of autism, in order to derive a dichotomous indicative autism variable. We did not consider any diagnoses to be mutually exclusive.

The CAPA was also used to establish the presence or absence of chronic irritability. A dichotomous irritability construct was generated, derived from the irritable dimension of oppositional defiant disorder (ODD); for this reason we chose not investigate the association between irritability and ODD, as the association would be likely to be inflated. The irritability items were selected on the basis of previous research, whereby the same items from the CAPA were used to derive an irritability construct^14,18^. The individual CAPA irritability items were coded as Absent (0) or present (1) and the items included were: 1. Temper tantrums 2. Touchy or easily annoyed 3. Angry and resentful. For this work, individuals were coded as irritable if the frequency of these irritable symptoms was chronic; ie. where one or more of these symptoms was present, 3–4 times/week in the past three months, participants were categorised as being “irritable” (1). Individuals who were coded as having an absence of all three of these symptoms, or the presence of these symptoms at frequencies lower than stated, were categorised as not irritable (0). We will refer to this chronic irritability presentation as “irritability” throughout (see table 1).

**TABLE 1:**
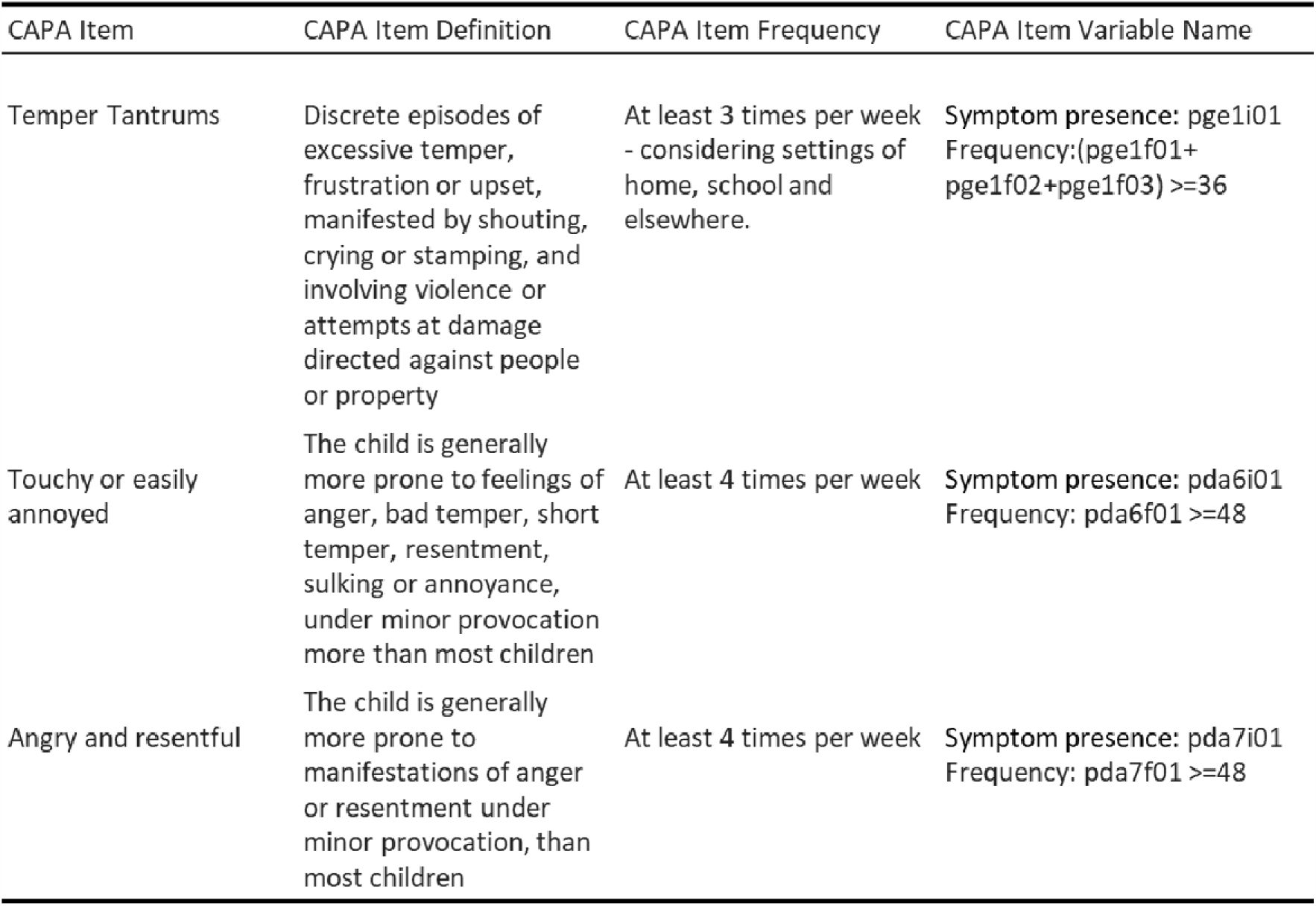
CAPA ITEMS INCLUDED IN THE IRRITABILITY CONSTRUCT. INDIVIDUALS WERE CODED AS IRRITABLE IF THEY HAD ONE OR MORE OF THE SYMPTOMS “TEMPER TANTRUMS”, “TOUCHY OR EASILY ANNOYED”, “ANGRY AND RESENTFUL”. ONE OR MORE OF THESE SYMPTOMS HAD TO BE PRESENT AT A FREQUENCY OF AT LEAST 3-4 TIMES PER WEEK, OVER THE LAST THREE MONTHS.

| CAPA Item | CAPA Item Definition | CAPA Item Frequency | CAPA Item Variable Name |
| --- | --- | --- | --- |
| Temper Tantrums | Discrete episodes of excessive temper, frustration or upset, manifested by shouting, crying or stamping, and involving violence or attempts at damage directed against people or property | At least 3 times per week - considering settings of home, school and elsewhere. | Symptom presence: pge1i01<br>Frequency: (pge1f01+pge1f02+pge1f03) >=36 |
| Touchy or easily annoyed | The child is generally more prone to feelings of anger, bad temper, short temper, resentment, sulking or annoyance, under minor provocation more than most children | At least 4 times per week | Symptom presence: pda6i01<br>Frequency: pda6f01 >=48 |
| Angry and resentful | The child is generally more prone to manifestations of anger or resentment under minor provocation, than most children | At least 4 times per week | Symptom presence: pda7i01<br>Frequency: pda7f01 >=48 |

#### Cognition

The young person’s intelligence quotient (IQ) was assessed using the Wechsler Abbreviated Scale of Intelligence (WASI), from which full scale IQ (FSIQ), verbal IQ (VIQ), and performance IQ (PIQ) composite scores are derived. The WASI includes four subtests; two performance subtests: non-verbal reasoning, perceptual organisation; and two verbal subtests: verbal knowledge and verbal reasoning.

### Analysis

The data were analysed using R version 4.0.1^25^ . Intraclass correlation (ICC) was carried out in IBM SPSS Statistics 27^26^. Odds ratios (ORs) and 95% confidence intervals (CIs) were derived. We included sibship as a random effect in all mixed effects models, to take into account the shared family factors for sibling pairs.

#### Aim 1

The first aim was to investigate whether young people with ND-CNVs are more irritable compared to controls. The percentage of young people with an ND-CNV and controls meeting criteria for irritability, and each of the irritability symptoms was calculated. We used a mixed effects logistic regression model, to investigate if age, sex, and ND-CNV status were associated with irritability.

#### Aim 2

To ascertain whether belonging to the same family (shared genetic and environmental factors) had an effect on irritability, an ICC was carried out in the subgroup of 123 available sibling pairs with one child with an ND-CNV and one with no known ND-CNV. ICC estimates were calculated based on a single-rating, absolute agreement, 2-way mixed effects model^27^. Intraclass correlation values of less than 0.5 are indicative of poor correlation, values between 0.5 and 0.75 indicate moderate correlation, values between 0.75 and 9 indicate good correlation, and values greater than 0.9 indicate excellent correlation^27^.

#### Aim 3

The next aim was to explore to what extent ND-CNV status is associated with irritability when controlling for other psychiatric diagnoses. We used mixed effects logistic regression to investigate if CNV-status was associated with irritability, controlling for age, sex and each of the following psychiatric diagnoses individually; ADHD, anxiety, indicative autism and mood disorder. Additionally, we used a mixed effects logistic regression model to investigate whether CNV-status was associated with irritability, when including a binary “any psychiatric diagnosis” variable, alongside age and sex, to account for the fact that individuals may have two or more comorbid psychiatric diagnoses.

Sensitivity analyses were conducted whereby mothers’ ethnicity, approximate family income and maternal education were included as covariates one by one alongside “any psychiatric diagnosis”, age and sex, for a subset of individuals where this information was available. Further sensitivity analyses were conducted, excluding those individuals who were taking psychiatric or epilepsy medication to establish if this would change the findings. Mixed effects binomial logistic regressions were also carried out with ADHD, anxiety, indicative autism, mood disorders and “any psychiatric diagnosis” as the outcome variables, with ND-CNV as the predictor variable, whilst controlling for irritability.

#### Aim 4

Finally, we aimed to study to what extent ND-CNV status is associated with irritability when controlling for cognitive ability in young people. A mixed effects logistic regression model, which included Full Scale IQ (FSIQ), Performance IQ (PIQ) and Verbal IQ (VIQ) one by one, alongside age and sex, was carried out. A separate logistic regression was carried out with the separate irritability items (Temper tantrums, Touchy or easily annoyed, Angry and resentful) as the outcome variable, and FSIQ as the predictor, specifically in ND-CNV carriers to test for within group differences.

## Results

### Aim 1 -Comparing irritability in young people with ND-CNV’s and controls

The prevalence of irritability was significantly greater in young people with an ND-CNV (54%), than controls (20%) (table 2). Irritability symptoms were also significantly higher in ND-CNV carriers than controls (see supplementary table 3), and of the young people with ND-CNVs there were an additional 119 (24.5%) young people with at least one irritable symptom, but at subthreshold frequency, compared with 30 (18.3%) controls. In line with previous research^6^ more young people with ND-CNVs met diagnostic criteria for psychiatric diagnoses; ADHD, anxiety and indicative autism, but not mood disorder (see table 2).

**TABLE 2:** DESCRIPTIVE AND SUMMARY STATISTICS FOR THE SAMPLE OF YOUNG PEOPLE WITH NEURODEVELOPMENTAL COPY NUMBER VARIANTS, AND CONTROLS.

| FAMILY BACKGROUND |  |  |  |
| --- | --- | --- | --- |
| Maternal Education | n = 569 |  |  |
| Low (O-levels, GCSEs) | 138 (24.3%) |  |  |
| Middle (A-levels, Highers, Vocational Training) | 240 (42.2%) |  |  |
| High (University/Post-graduate degree) | 142 (24.9%) |  |  |
| No school leaving exams | 49 (8.6%) |  |  |
| Approximate Family Income | n = 520 |  |  |
| < = £ 19 999 | 150 (28.8%) |  |  |
| £ 20 000 - £ 39 999 | 180 (34.6%) |  |  |
| £ 40 000 - £ 59 999 | 97 (18.7%) |  |  |
| £ 60 000 + | 93 (17.9%) |  |  |
| Mothers Ethnicity | n = 579 |  |  |
| European | 536 (92.6%) |  |  |
| Other | 43 (7.4%) |  |  |
| DEMOGRAPHICS OF YOUNG PEOPLE |  |  |  |
|  | ND-CNV | Controls | Statistics |
| Age | n= 485 | n=164 |  |
| Mean | 9.8 | 10.8 | *p* < 0.001 |
| Biological Sex | n= 485 | n=164 |  |
| Female | 168 (34.6%) | 77 (47.0%) | *p* = 0.005 |
| Male | 317 (65.4%) | 87 (53.0%) |  |
| PSYCHIATRIC DIAGNOSES |  |  |  |
|  | n= 485 | n=164 |  |
| ADHD | 225 (46.4%) | 12 (7.3%) | *p* < 0.001 |
| Anxiety | 163 (33.6%) | 11 (6.7%) | *p* < 0.001 |
| Mood | 12 (2.5%) | 1 (0.6%) | *p* = 0.141 |
| Any Psychiatric Diagnosis | 348 (71.8%) | 24 (14.7%) | *p*<0.001 |
| AUTISM |  |  |  |
|  | n=409 | n=145 |  |
| SCQ >15 | 237 (57.9%) | 8 (5.5%) | *p*<0.001 |
| COGNITION |  |  |  |
|  | n= 433 | n= 159 |  |
| Mean FSIQ | 80.4 | 104.8 | *p* < 0.001 |
| Mean PIQ | 86.3 | 105.1 | *p* < 0.001 |
| Mean VIQ | 80.2 | 104.1 | *p* < 0.001 |
| IRRITABILITY |  |  |  |
|  | n= 485 | n=164 |  |
| % Irritable | 54.2 | 20.2 | *p* < 0.001 |

A mixed effects binomial logistic regression, showed that irritability was associated with ND-CNV-status (OR = 3.85, CI = 3.00-7.71, p= 1.02 x 10^-10^) with young people with ND-CNVs more likely to be irritable. Irritability was also associated with sex (OR = 1.48, CI = 1.05 – 2.15, p=0.02), with males more likely to meet irritability criteria. There is also a significant interaction effect of ND-CNV*Sex (OR=2.77, CI = 1.13 - 6.84, p = 0.03), thus males with a ND-CNV are more likely to be irritable than females with a ND-CNV. Age was not associated with the presence of irritability.

### Aim 2 - To what extent do shared genetic and environmental factors in families predict irritability ?

We compared the degree to which irritability in young people with a ND-CNV could be predicted by irritability in their siblings, to evaluate possible contributions of shared genetic and environmental factors. A low degree of correlation in irritability was found between young people with ND-CNVs and their siblings with no known ND-CNV. The single measure ICC was 0.066 (CI = -0.079 - 0.221, p = 0.183), suggesting that shared familial effects have little effect on the irritability outcome.

### Aim 3 - To what extent is ND-CNV status associated with irritability when controlling for other psychiatric diagnoses?

A series of mixed effects binomial logistic regressions testing in turn ADHD, anxiety, indicative autism and mood disorder showed that irritability was associated with a diagnosis of ADHD (OR = 2.72, CI = 1.95-4.18, p = 7.67×10^-8^), anxiety (OR = 3.49, CI = 2.65-6.67, p= 1.35×10^-9^) and indicative autism (OR =2.41, CI = 1.60-3.73, p = 3.53×10^-5^). Irritability showed borderline association with a diagnosis of mood disorder (OR = 1.45, CI = 1.03-18.57, p = 0.06). Age was not associated with the presence of irritability when controlling for psychiatric diagnoses. Sex was not associated with irritability when controlling for ADHD or indicative autism, however, sex was associated with irritability when controlling for anxiety (OR = 1.48, CI = 1.03 - 2.18, p = 0.03) and mood disorder (OR = 1.46, CI = 1.04 - 2.13, p = 0.03), with males more likely to be irritable.

CNV status was still associated with irritability, when controlling for ADHD (OR = 2.75, CI =2.17-5.75, p= 1.47×10^-6^), anxiety (OR = 3.49, CI = 2.65-6.67, p =5.22×10^-7^), indicative autism (OR = 2.55, CI = 1.71 - 5.08, p = 1.26×10^-4^) and mood disorder (OR = 3.78, CI =1.03-7.56, p = 2.01×10^-10^). This remained the case after excluding individuals taking psychiatric or epilepsy medication.

To allow for the fact that individuals may have one or more comorbid psychiatric conditions, a dichotomous variable of “any psychiatric diagnosis”, was included in a mixed effects logistic regression alongside age and sex, with irritability as the dependent variable. “Any psychiatric diagnosis”, was also associated with irritability (OR = 3.20, CI = 2.17 - 4.50, p =2.52×10^-8^). CNV-status was still associated with irritability when controlling for “any psychiatric diagnosis” (OR = 2.22, CI =1.54-4.21, p = 3.26×10^-4^). Age and sex were not associated with irritability when controlling for any psychiatric diagnosis. This remained the case after excluding individuals taking psychiatric or epilepsy medication.

Sensitivity analyses including maternal education, approximate family income, and mothers ethnicity as covariates, alongside “any psychiatric diagnosis”, age and sex, found that mothers’ education, mothers ethnicity, and approximate family income were not significantly associated with irritability. Including these variables as covariates in the analysis did not change the findings.

Mixed effects binomial logistic regressions were also carried out with ADHD, anxiety, indicative autism, mood disorders and “any psychiatric diagnosis” as the outcome variables, with ND-CNV as the predictor variable, whilst controlling for irritability. The associations between ND-CNVs and psychiatric diagnoses remained significant (see supplementary table 4).

### *Aim 4:* To what extent does cognition explain ND-CNV associations with irritability?

In line with previous research, young people with ND-CNVs in this study had significantly lower full scale IQ, performance IQ, and verbal IQ scores (table 1), than controls. Young people with ND-CNVs had on average a 24.4 point lower FSIQ score, an 18.8 point lower PIQ score and a 24.8 point lower VIQ compared with controls. A mixed effects logistic regression testing in turn FSIQ, PIQ and VIQ whilst co-varying for age and sex found no association between irritability and FSIQ (OR=1.02, CI=0.99-1.03, p=0.93), PIQ (OR=0.98, CI=0.80-1.01, p=0.40) or VIQ (OR=0.97, CI=0.88-1.02, p=0.89). ND-CNV status was still associated with irritability when controlling for FSIQ (OR=4.08, CI =2.91-7.72, p =6.98×10^-8^), PIQ (OR=3.86, CI=2.90-7.98, p=2.85×10^-9^) and VIQ (OR =3.96, CI =2.99-7.81, p =6.33×10^-8^). This remained the case after excluding individuals taking psychiatric or epilepsy medication. No significant associations were found between FSIQ and the three irritability symptoms in those with ND-CNVs (supplementary table 5).

## Discussion

### Main findings

This is the first, and largest study, as far as we are aware, that investigates irritability of a chronic nature in young people with a range of recurrent CNVs. CNVs are increasingly diagnosed by Medical Geneticists and well documented to be associated with high risk of neurodevelopmental and psychiatric conditions (ND-CNVs)^6^. Irritability was established based on items from a semi-structured child and adolescent psychiatric interview (CAPA)^23^ completed with the primary carer. This is an irritability construct which has been used previously in other work pertaining to irritability, and neurodevelopmental disorders^14,15,18^. Young people were considered irritable if they demonstrated temper tantrums, angry and resentful behaviour or were touchy or easily annoyed, 3-4 times per week in the last three months^12^. We demonstrate that young people with ND-CNVs are close to 4 times (OR=3.85, CI= 3.00-7.71, p=1.02 x 10^-10^) more likely to show irritability than controls. Irritability was present in 54% of young people with ND-CNVs compared with 20% of controls. Irritability was associated with sex and CNV status, with males and those with ND-CNVs more likely to meet the threshold for severe irritability. Our results did not indicate that mothers’ education, ethnicity, or approximate family income affected irritability. Irritability was not associated with cognition (FSIQ, PIQ or VIQ).

Our findings offer supporting evidence for previous studies which have described high rates of psychiatric and neurodevelopmental difficulties, and cognitive deficits in individuals with ND-CNVs^4,6,8^. We found no differences in mood disorders between young people with ND-CNVs and controls. The numbers of young people in the sample who met criteria for mood disorders was low, and this is likely due to the age of the sample. We found that irritability in young people with ND-CNVs was associated with a diagnosis of ADHD, anxiety, and indicative autism. Our findings are also in line with research which suggests that irritability is a common feature of ADHD^14,15^, anxiety^16^ and autism^10,11,19^.

When controlling for psychiatric diagnoses, ND-CNV status remained a significant predictor of irritability. This demonstrates that whilst individuals with ND-CNVs are likely to have comorbid psychiatric diagnoses which are associated with increased irritability, the impact of the ND-CNV on irritability is at least partially independent of the association between ND-CNVs and psychiatric diagnoses of ADHD, anxiety, indicative autism and mood disorder. When controlling for irritability, the association between ND-CNVs and psychiatric disorders remains significant, suggesting that the association between ND-CNVs and psychiatric disorders are at least to some extent, independent of the association between ND-CNV and irritability. Given that individuals with ND-CNVs are at high risk for depression and schizophrenia^8^, and irritability can be a feature of both diagnoses^11^, the early presentation of irritability may be an important transdiagnostic phenotype in these individuals, and further work investigating longitudinal trajectories would be pertinent to potentially inform early intervention.

Additionally, whilst young people with ND-CNVs have significantly lower IQ scores, these were not associated with irritability. This is in line with studies in children with ADHD, which have found little evidence of links between irritability and cognitive functioning^28^. ND-CNV status was still associated with irritability when controlling for cognitive ability, thus, the association between ND-CNVs and irritability appears to be independent of the association between ND-CNVs and reduced cognitive ability.

We found that males in general were more likely to meet criteria for irritability than females. Sex differences in irritability are seemingly complex, and previous findings suggest that male preponderance may be a feature of neurodevelopmental problems whereas female preponderance is a feature of mood and internalising problems^15,29^. Longitudinal follow up will help to ascertain if this association between sex and irritability is stable or dynamic in young people with ND-CNVs.

It should be acknowledged that whilst young people with ND-CNVs have higher rates of irritability than controls, 20% of the sibling control group met criteria for irritability. Findings drawn from the Great Smoky Mountains study suggests that severe mood dysregulation (in which chronic irritability is present) occurs in children and adolescents in approximately 3% of the general population, and that at any given point in childhood, 51.4% of children report phasic irritability^19^. Further investigation would be required to determine a) whether siblings of ND-CNV groups have significantly higher rates of chronic irritability than the general population, when controlling for age and sex. There was little correlation in irritability between the young people with ND-CNVs and their siblings; the siblings of young people with ND-CNVs with irritability do not have higher rates of irritability than the siblings of young people with ND-CNVs who are not irritable. In line with previous findings^30^, this suggests that shared environmental and genetic factors have a relatively small effect on irritability.

With regards to treatment for irritability, research suggests that parent management training (PMT) and cognitive behavioural therapy (CBT) are the most effective treatments^31^. However, given the comorbidity of irritability with other psychiatric conditions in young people with ND-CNVs, consideration should be given to the type of intervention which will be most effective. For instance Stepping Stones Triple P has shown efficacy as a positive parenting intervention for parents of children with learning disabilities^32^. Additionally, although low IQ does not predict irritability in young people with ND-CNVs, young people with ND-CNVs who are irritable may well have low IQ, and adaptations to CBT should be considered in these instances^33^.

### Limitations

Some caveats to this study should be noted. Firstly, ascertainment bias may affect our results, that is, our cohort of young people with ND-CNVs is likely to be more severely affected than young people with ND-CNVs recruited as part of a population-based cohort. This is because we recruited individuals with a known genetic diagnosis, where referral for genetic testing is often based on the presence of developmental delay. There is however increasing evidence that CNVs impact on functioning in the general population including in individuals without a genetic diagnosis^4^. Furthermore, it is important to understand irritability in those diagnosed with a ND-CNV, as they constitute a substantial proportion of those engaging with health services and tend to need more support and care following their genetic diagnosis. Secondly, due to the level of comorbidity in the sample, irritability cannot be examined in the absence of comorbid psychiatric diagnoses; this is however, reflective of the reality outside of the exploration of young people with ND-CNVs, as irritability is such a common feature of so many psychiatric diagnoses. Finally, this study does not capture all the environmental factors which may influence irritability in young people with ND-CNVs.

### Implications

Irritability is an important aspect of the clinical picture in young people with recurrent ND-CNVs and cannot simply be attributed to IQ impairment, or the presence of other psychiatric diagnoses. Clinicians thus need to be aware of the high prevalence of irritability in this group of high-risk individuals. This finding also gives new insights into the genetic aetiology of irritability. The frequent co-occurrence of irritability with ADHD, anxiety and indicative autism suggests that clinicians working with young people with ND-CNVs should assess comorbid symptom profiles, in order to provide optimum care and the correct intervention. Future research should investigate irritability longitudinally, to ascertain whether it may be an early marker for later psychiatric problems such as mood disorders and psychosis. On the whole, irritability warrants attention as a feature in young people with ND-CNVs, in order to inform interventions for irritable behaviour and comorbid psychopathologies.

## Data Availability

Data are held at Cardiff University. The data are available upon request. Please contact Prof. van den Bree for any data requests.

## Acknowledgements

We are extremely grateful to all the families that participated in this study, as well as all the support we have had from NHS medical genetics clinics, and from support charities; Max Appeal, Unique and the 22Crew. We thank all members of the IMAGINE-ID consortium for their contributions (see supplementary material for full consortium member list). We would also like to thank the following people for contributing to data collection; Samantha Bowen, Molly Tong, Philippa Birch, Matthew Sopp, Alice Robinson, Sinéad Ray, Nicola Lewis, Sarah Law, Sophie Andrews, Aimée Challenger, Charlotte Thomas-Carmichael, Lucy Reed, Lauren Benger, Megan Thomas, Poppy Sloane, Alice Walsh, Keziah Fish, Amy Ilsley, Stephen Naughton, Rachel Tompkins, Ciara Walker, Nadia Pantouw, Hannah Pendlebury, Chloe Sheldon, Emily Green, Umaya Prasad, Joshua Roberts, Jessica Townsend and Beth Hughes. We thank Hayley Moss for research management support and Karen Bradley for administrative support. We thank the core laboratory team of the Division of Psychological Medicine and Clinical Neurosciences laboratory for DNA sample management and genotyping. We also thank the National Centre for Mental Health, a collaboration between Cardiff, Swansea and Bangor Universities, for their support. For the purpose of Open Access, the author has applied a CC BY public copyright license to any Author Accepted Manuscript (AAM) version arising from this submission.

