## Supplementary material for "Irritability in young people with copy number variants associated with neurodevelopmental disorders (ND-CNVs)"

**Supplementary Information**

| **Genotype** | **N** | **Critical Region (hg19)** |
| --- | --- | --- |
| Controls | 164 |  |
| 22q11.2 Deletion | 150 | chr22:19,037,332-21,466,726 |
| 16p11.2 deletion | 65 | chr16: 29,650,840-30,200,773 |
| 15q11.2 deletion | 47 | chr15:22,805,313-23,094,530 |
| 1q21.1 duplication | 29 | chr1:146,527,987-147,394,444 |
| 16p11.2 duplication | 28 | chr16: 29,650,840-30,200,773 |
| 15q13.3 deletion | 27 | chr15:31,080,645-32,462,776 |
| 22q11.2 duplication | 27 | chr22:19,037,332-21,466,726 |
| 1q21.1 deletion | 25 | chr1:146,527,987-147,394,444 |
| More than one ND-CNV | 22 |  |
| 2p16.3 (NRXN1) deletion | 20 | chr2:50145643-51259674 |
| 15q13.3 duplication | 17 | chr15:31,080,645-32,462,776 |
| 16p11.2 distal deletion | 15 | chr16:28,823,196-29,046,783 |
| 1q21.1 TAR duplication | 13 | chr1:145,394,955-145,807,817 |
| **TOTAL** | **649** |  |

**supplementary table 1: description of number of participants with each genotype within sample**

| **Medication** | **Number of Individuals** |
| --- | --- |
| Adderall | 1 |
| Aripiprazole | 1 |
| Atomoxetine | 4 |
| Carbamazepine | 1 |
| Fluoxetine | 3 |
| Lamotrigine | 2 |
| Lithium | 1 |
| Methylphenidate | 5 |
| Risperidone | 1 |
| Sertraline | 3 |
| Sodium Valproate | 4 |
| Topiramate | 1 |

**supplementary table 2: psychiatric and epilepsy medications being used in sample. adderall, atomoxetine, methylphenidate, are typically prescribed to treat adhd. carbamazepine, lamotrigine, sodium valproate and topiramate are typically prescribed to treat epilepsy. risperidone and aripiprazole are antipsychotic medications which may have also been prescribed “off label” to treat adhd, autism, obsessive compulsive disorder, depression, behavioural disorders, tics or tourettes. lithium is used to treat bipolar disorder. sertraline and fluoxetine are selective serotonin reuptake inhibitors typically used to treat depression and anxiety.**

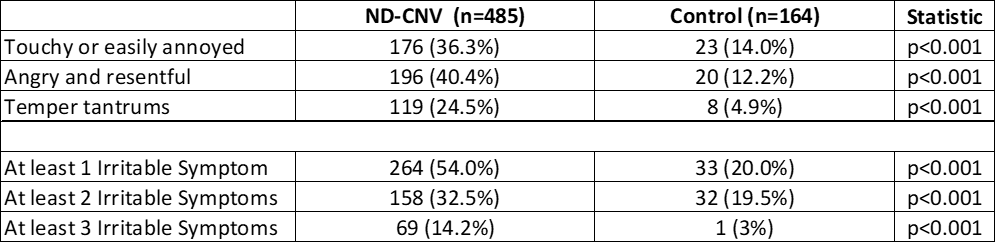

**Supplementary table 3: number and proportion of nd-cnv carriers and controls who met criteria for each of the symptoms in the irritability construct; touchy or easily annoyed, angry and resentful, and temper tantrums. To meet criteria for each symptom, the symptom had to be present at least 3-4 times per week in the last three months. Table also presents the number and proportion of individuals who met for at least 1, 2 or 3 of the symptoms from the irritability construct.**

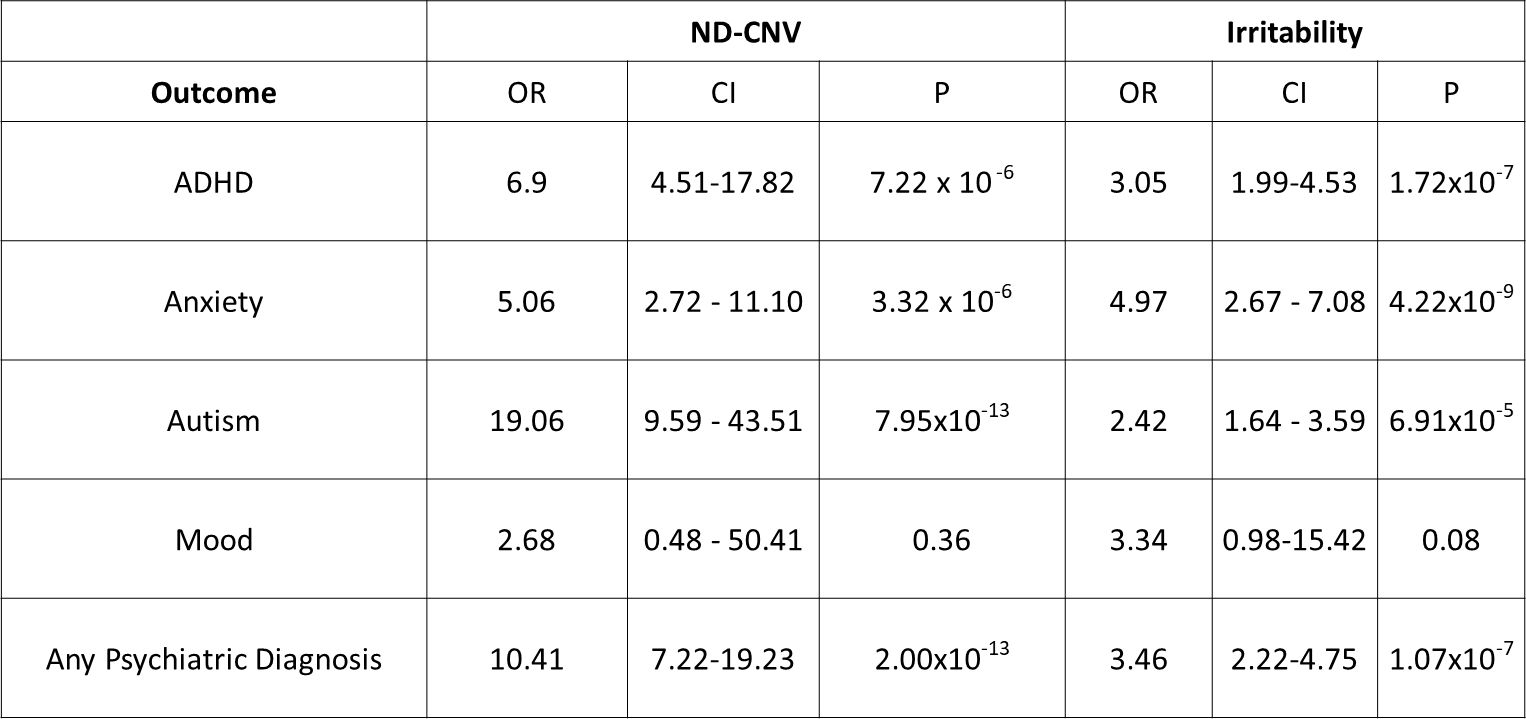

**supplementary table 4: results from mixed effects logistic regression models with adhd, anxiety, indicative autism, mood disorders and “any psychiatric diagnosis” as the outcome variables with nd-cnv as the predictor variable, controlling for irritability, age and gender. Nd-cnv status and irritability are associated with adhd, anxiety, indicative autism and any psychiatric diagnosis, but not mood. This demonstrates that the association of nd-cnvs with psychiatric disorders is, at least in part, independent of the association between nd-cnvs and irritability.**

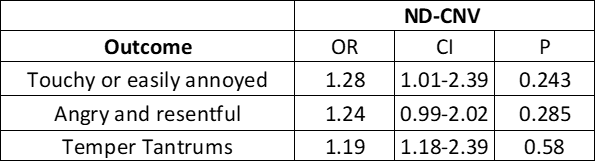

**Supplementary table 5: results from logistic regression in nd-cnv carriers, with each of the three irritable symptoms as the outcome variable, and fsiq as the predictor, controlling for age and gender. There is no association between fsiq and each of the irritable symptoms in nd-cnv carriers.**

**IMAGINE-ID consortium members**

**IMAGINE-ID Consortium author list**

| **Surname** | **Initials** | **First Name** | **Title** | **Institution** |
| --- | --- | --- | --- | --- |
| **Raymond** | F L | F Lucy | Professor | Department of Medical Genetics, University of Cambridge, UK |
| **Dewhurst** | E | Eleanor | Mrs | Department of Medical Genetics, University of Cambridge, UK |
| **Lafont** | A | Amy | Ms | Department of Medical Genetics, University of Cambridge, UK |
| **Timur** | H | Husniye | Ms | Department of Medical Genetics, University of Cambridge, UK |
| **Wicks** | F | Francesca | Mrs | Department of Medical Genetics, University of Cambridge, UK |
| **Ye** | Z | Zheng | Dr | Department of Medical Genetics, University of Cambridge, UK |
| **Baker** | K | Kate | Dr | Department of Medical Genetics, University of Cambridge, UK |
| **Walker** | N | Neil | Dr | Department of Medical Genetics, University of Cambridge, UK |
| **Wallwork** | S | Sarah | Ms | Department of Medical Genetics, University of Cambridge, UK |
| **Skuse** | D | David | Professor | Great Ormond Street Institute of Child Health, University College London, UK |
| **Denaxas** | S | Spiros | Dr | Institute of Health Informatics, University College London, London, UK |
| **Mandy** | W | William | Dr | Division of Psychology & Language Sciences, University College London, UK |
| **Wolstencroft** | J | Jeanne | Dr | Great Ormond Street Institute of Child Health, University College London, UK |
| **Davies** | S | Sarah | Ms | Great Ormond Street Institute of Child Health, University College London, UK |
| **Erwood** | M | Marie | Ms | Great Ormond Street Institute of Child Health, University College London, UK |
| **Juj** | M | Manoj | Mr | Great Ormond Street Institute of Child Health, University College London, UK |
| **Kerry** | E | Eleanor | Ms | Great Ormond Street Institute of Child Health, University College London, UK |
| **Lucock** | A | Anna | Ms | Great Ormond Street Institute of Child Health, University College London, UK |
| **Printzlau** | F | Frida | Ms | Great Ormond Street Institute of Child Health, University College London, UK |
| **Srinivasan** | R | Ramya | Dr | Great Ormond Street Institute of Child Health, University College London, UK |
| **Walker** | S | Susan | Dr | Great Ormond Street Institute of Child Health, University College London, UK |
| **Coscini** | N | Nadia | Dr | Great Ormond Street Institute of Child Health, University College London, UK |
| **Fatih** | N | Nasrtullah | Mr | Great Ormond Street Institute of Child Health, University College London, UK |
| **Denyer** | H | Hayley | Ms | Great Ormond Street Institute of Child Health, University College London, UK |
| **Andrews** | S | Sophie | Ms | MRC Centre for Neuropsychiatric Genetics and Genomics, Division of Psychological Medicine and Clinical Neurosciences, Cardiff University, UK |
| **Chawner** | SJRA | Samuel | Dr | MRC Centre for Neuropsychiatric Genetics and Genomics, Division of Psychological Medicine and Clinical Neurosciences, Cardiff University, UK |
| **Cuthbert** | A | Andrew | Dr | MRC Centre for Neuropsychiatric Genetics and Genomics, Division of Psychological Medicine and Clinical Neurosciences, Cardiff University, UK |
| **Challenger** | A | Aimee | Ms | MRC Centre for Neuropsychiatric Genetics and Genomics, Division of Psychological Medicine and Clinical Neurosciences, Cardiff University, UK |
| **Hall** | J | Jeremy | Professor | MRC Centre for Neuropsychiatric Genetics and Genomics, Division of Psychological Medicine and Clinical Neurosciences, Cardiff University, UK |
| **Lewis** | N | Nicola | Ms | MRC Centre for Neuropsychiatric Genetics and Genomics, Division of Psychological Medicine and Clinical Neurosciences, Cardiff University, UK |
| **Owen** | MJ | Michael | Professor Sir | MRC Centre for Neuropsychiatric Genetics and Genomics, Division of Psychological Medicine and Clinical Neurosciences, Cardiff University, UK |
| **Ray** | S | Sinead | Ms | MRC Centre for Neuropsychiatric Genetics and Genomics, Division of Psychological Medicine and Clinical Neurosciences, Cardiff University, UK |
| **Sopp** | M | Matthew | Mr | MRC Centre for Neuropsychiatric Genetics and Genomics, Division of Psychological Medicine and Clinical Neurosciences, Cardiff University, UK |
| **Moss** | H | Hayley | Ms | MRC Centre for Neuropsychiatric Genetics and Genomics, Division of Psychological Medicine and Clinical Neurosciences, Cardiff University, UK |
| **van den Bree** | MBM | Marianne | Professor | MRC Centre for Neuropsychiatric Genetics and Genomics, Division of Psychological Medicine and Clinical Neurosciences, Cardiff University, UK |
| **Holmans** | P | Peter | Professor | MRC Centre for Neuropsychiatric Genetics and Genomics, Division of Psychological Medicine and Clinical Neurosciences, Cardiff University, UK |
| **Bowen** | S | Samantha | Ms | MRC Centre for Neuropsychiatric Genetics and Genomics, Division of Psychological Medicine and Clinical Neurosciences, Cardiff University, UK |
| **Bradley** | K | Karen | Mrs | MRC Centre for Neuropsychiatric Genetics and Genomics, Division of Psychological Medicine and Clinical Neurosciences, Cardiff University, UK |
| **Birch** | B | Philippa | Ms | MRC Centre for Neuropsychiatric Genetics and Genomics, Division of Psychological Medicine and Clinical Neurosciences, Cardiff University, UK |
| **Tong** | M | Molly | Ms | MRC Centre for Neuropsychiatric Genetics and Genomics, Division of Psychological Medicine and Clinical Neurosciences, Cardiff University, UK |
| **Ford** | T | Tasmin | Professor | Department of Psychiatry, University of Cambridge |
| **Searle** | B | Beverly | Dr | Unique Charity, UK |
| **Wynn** | S | Sarah | Dr | Unique Charity, UK |
| **Robertson** | L | Lisa | Dr | Aberdeen Royal Infirmary Genetics Service |
| **Berg** | J | Jonathan | Dr | Ninewells Hospital Dundee Genetics Service |
| **Lampe** | A | Anne | Professor | Western General Hospital Edinburgh Genetics Service |
| **Joss** | S | Shelagh | Dr | Glasgow Genetics Centre, Glasgow |
| **Brennan** | P | Paul | Dr | Northern Genetics Service, Newcastle |
| **Kraus** | A | Alison | Dr | Yorkshire Regional Genetics Service - Clinical Genetics |
| **Weber** | A | Astrid | Dr | Cheshire & Merseyside Regional Genetic Service |
| **Rawson** | M | Myfanwy | Ms | Manchester Centre for Genomic Medicine |
| **Quarrell** | O | Oliver | Dr | Sheffield Genetic Services |
| **Vasudevan** | P | Pradeep | Dr | Leicestershire Genetics Centre, Leicester |
| **Harrison** | R | Rachel | Dr | Nottingham Regional Genetics Service |
| **Williams** | D | Denise | Dr | West Midlands Regional Genetics Service, Birmingham |
| **Maher** | E | Eamonn | Professor | East Anglian Medical Genetics Service, Cambridge |
| **Kini** | U | Usha | Dr | Oxford Genetics Service |
| **Clowes** | V | Virginia | Dr | London North West Thames Regional Genetics Service |
| **Van Dijk** | F | Fleur | Dr | London North West Thames Regional Genetics Service |
| **Gurasashvilli** | J | Jana | Dr | London North East Thames Regional Genetics Service - Great Ormond Street Hospital, London |
| **Mansour** | S | Sahar | Dr | London South West Thames Regional Genetics Service, St Georges Hospital, Tooting, London |
| **Holder-Espinasse** | M | Muriel | Dr | London South East Thames Regional Genetics Service Guy's Hospital, London |
| **Watford** | A | Amy | Dr | Bristol Clinical Genetics Service, Bristol |
| **Rankin** | J | Julia | Dr | Peninsula Genetics Service, Exeter |
| **Baralle** | D | Diana | Dr | Wessex Clinical Genetics Service |
| **Procter** | A | Annie | Dr | All Wales Regional Genetics Service |
